# Deep Stroop: Using eye tracking and speech processing to characterize people with neurodegenerative disorders while performing the Stroop Test

**DOI:** 10.1101/2023.05.30.23290742

**Authors:** Trevor Meyer, Anna Favaro, Tianyu Cao, Ankur Butala, Esther Oh, Chelsie Motley, Pedro Irazoqui, Najim Dehak, Laureano Moro-Velázquez

## Abstract

Although many neurodegenerative diseases affect different neural circuits, they often express complex and overlapping symptom profiles making them difficult to differentiate precisely. Current methods of analyzing patients are limited to bedside examination, patient self-rating scales, semiquantitative clinician-rating scales, and other observational evidence, which are often non-specific, resulting in open multiple interpretations and ambiguity in diagnosis and treatment plans. We present a method to analyze patient symptom profiles using multimodal analysis of subjects performing the Stroop Test. We use high-sample-rate eye tracking and speech recording tools to record subject behavior while completing the Stroop Test and simultaneously analyze multiple traits of their interaction with the test. We compare the performance of healthy controls to patients with Parkinson’s Disease, Alzheimer’s Disease, and other neurodegenerative diseases with clinical parkinsonism. We automatically extract metrics based on eye motor behavior, gaze characteristic uttered responses, and the temporal relationship between gaze and uttered responses. We identify many that have clinical relevance through high correlations with existing MoCA and MDS-UPDRS, many of which have significantly different distributions between groups. We present here our analysis approach, provide freely available source code to replicate it and demonstrate the potential of multi-modal recording and analysis of patients throughout their execution of neuro-psychological tests like the Stroop Test.

## 1. Introduction

Neurodegenerative Diseases (NDs) affect millions of people worldwide, and with an aging population, we will surely face an increasing burden. Nearly 1 million people in the US are living with Parkinson’s Disease (PD), with 60,000 new diagnoses each year [1], and as many as 6.2 million Americans may have Alzheimer’s Disease (AD) [2]. PD and AD are among the most common, yet NDs include a wide range of chronic conditions that result from the damage or loss of function of cells in the nervous system and often manifest overlapping symptom profiles. While there exist lots of high-quality resources to analyze specific dysfunctions indicative of specific NDs [3, 4, 5, 6], there has been a lack of progress in accumulating all of these observations into a comprehensive analytic and diagnostic tool with clinical relevance. Much of the analytical and diagnostic approaches in use today rely on a restricted number of evaluations by a specialist, anecdotal evidence from the patient or an informant, or other clinical rating scales that are often subjective, nonlinear, and limited in the depth of their insight into ND presence or progression. These tests are also rarely specific, leaving opportunities for multiple valid interpretations and diagnoses, leading to inconsistencies and errors in the treatment plans of neurological disease patients. Lastly, existing hub-and-spoke healthcare models accentuate accessibility disparities in which patients living around large academic medical centers readily access advanced diagnostic testing (i.e., functional imaging, genetic testing). There is a need for cost-efficient, easily deployable, reliable, comprehensive, quantitative tools which provide doctors and researchers with richer analysis and allow them to benchmark patient performance and track progress over time.

Fusing functional and cognitive modalities with neuropsychological tests presents a unique opportunity to characterize subject behavior and dysfunctions for deeper and more comprehensive analysis. In this study, we offer the combined analysis of eye tracking and speech recordings to analyze subjects with various NDs during the execution of the Stroop Test (ST). We use these two modalities to perform in-depth high-precision analysis of subjects throughout ST progression, generating high-resolution and detailed metrics. We employ this method to expand significantly on the analytical value of the ST in the context of ND detection and differentiation. While considering final output and whole-test performance provides some valuable information, subjects’ interaction with these tests and compensation strategies, if detectable, could provide even more insight into subject behavior and impairment, allowing for specific phenotyping and symptom tracking. We show the validity of these measurements in NDs through strong correlations with existing well-accepted motor and cognitive screening tests like UPDRS and MoCA and also propose new findings and metrics that may better represent and characterize patient phenotypes, allowing for detection and differentiation of PD, NDs with symptoms mimicking PD (PDM), AD, and Healthy Controls (CTL).

## 2. Background

### 2.1. Eye Movement

Analyzing eye movement in a testing environment involves the consideration of the four types of human eye motion states: saccade, fixation, smooth pursuit, and blink. Saccades involve the rapid movement of the eyes from one point to another, often lasting less than 100 *ms* [7] at a speed greater than 30 *deg/s* [8]. Saccades are understood to be simple ballistic movements; no visual information can be interpreted during a saccade due to the high rate of change of visual stimuli. Fixations involve resting the eye gaze on a single point in space for some duration in time, often greater than 100 *ms* [7]. During fixations, visual stimuli focus on the fovea and thus can be processed and interpreted by the visual system. Smooth Pursuit involves the tracking, or pursuit, of a stimulus of interest as it moves in space utilizing slow, steady eye movement. The stimulus of interest remains focused and can be processed and interpreted by the visual system, even as the eye moves. Lastly, a blink involves rapidly closing the eyelids over the eyes, briefly obstructing the visual system. These are necessary to protect, clean, and rehydrate the eye’s surface. While the eye typically does move during a blink [9], we do not consider that movement in this study.

Even a single saccade engages many neural circuits and brain areas to initiate and coordinate this highly precise directed motion, including the midbrain, cerebellum and extra-cerebellar nuclei, basal ganglia, thalamus, and multiple far-flung of the cerebral cortex. They can be voluntary or subconscious, are among some of the most well-studied and understood human motor responses [7], and are well suited for detecting and studying neurological degeneration. Dysfunction of different neural circuits at each stage of eye movement control can have predictable and detectable effects on resulting eye movement patterns and may be indicative of specific NDs. By identifying the sequence and characteristics of these states of eye movement, we can quantify and analyze behavioral patterns in subjects at rest and throughout the completion of tasks.

One emerging technology with great potential to capture this behavior with high levels of resolution and detail is high-sampling-rate eye tracking systems, which can estimate eye movement and gaze position. Modern systems can even be performed without contact, even on patients wearing personal glasses, allowing for observation of patients’ unobstructed behavior patterns as they interact with prompts and activities on a screen. In addition to analyzing a range of oculomotor abnormalities linked to NDs - which are described in detail below - eye tracking provides unique insights into the behavior and performance of subjects, acting as a new measure of information input to neural processing systems alongside the classically analyzed measures of the generated output.

### 2.2. Speech Production

Speech and language impairments are connected to motor and cognitive decline, emerging at varying stages across NDs. Speech production requires specific cognitive and functional brain areas to coordinate complex supra-glottal articulators, lungs, and larynx movements. These areas of the brain can be damaged by NDs in predictable ways, consequently influencing human communication skills as well as degenerating vocal performance [10], and can be valid predictors of ND onset and earlier indicators of its progression [11, 12, 13, 6].

Thus, detailed speech analysis can help identify patterns that cannot be easily distinguished from a clinical standpoint and thus help perform earlier diagnosis of NDs and monitor their progression [14]. The analysis of speech production can interest the phonetic and phonological components of vocal sound, lexico-semantic and morpho-syntactic components of word content, and pragmatic levels of language organization [14]. Many studies employ cognitive tasks (e.g., picture description, interview, reading passage) to elicit connected speech in NDs. These have different contributions to evaluating other phonatory, articulatory, and linguistic domains [14].

### 2.3. Stroop Test

Initially presented in 1935 by J. R. Stroop [15], the Stroop Test (ST), also called the word-color naming test, has been used extensively in psychological and neurological contexts to measure neural function and cognitive performance. For instance, it is currently used as a standard test in the Lewy-body Dementia Module of the US National Alzheimer’s Coordinating Center ^1^. The ST involves three phases of stimuli presentation, where performance is independently quantified in each task and later combined for scoring. A grid of text is presented to the subject in each task. First, the subject is instructed to utter aloud the word that is spelled in black text in task 1 (word-naming), then the color of displayed “####” characters in task 2 (color-naming), and finally the color of the text, not the word that is spelled, in task 3 (word-color-naming; incongruent or conflicting stimuli). The word-color-naming task is shown in Figure 1 and is the most difficult of the three tasks requiring the subject to inhibit the faster/more automatic response of reading the text (and instead must utter the text color.

**Figure 1:**
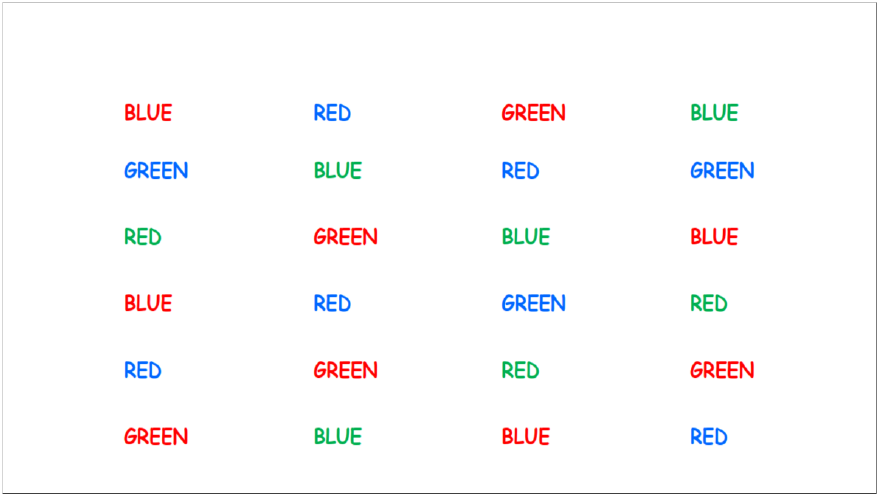
An example presentation of the word-color-naming task of the ST Subjects are instructed to say aloud the color of the text of each word, progressing from Left to Right and Top to Bottom. In this presentation, the correct answers of the first row would be “Red, Blue, Red, Green”

While there is no standardized format or scoring procedure for the ST, all accepted implementations generate three scores based on either the completion time (seconds) or progress (number of words) through color-naming, word-naming, and word-color-naming tasks of the test [16, 17]. While there are many valid methods to normalize and combine these scores for analysis, reviews of available methods suggest that three independent phenomena are measurable through classic scoring metrics: reading time, color-naming difficulty, and interference management [17]. These scores consistently reveal a pattern coined the “Stroop Effect”, which involves performing the best on word-naming, followed by color-naming, and performing worst on word-color-naming where the word-naming and color-naming responses interfere. There has also been some analysis of error rate and error-correction rate [18], but this has otherwise been the limit of the quantification of ST performance. Some studies suggest performance decline with increasing age [19, 20] and decreasing level of education, along with minor gender differences sometimes reported [20].

The ST has a long-established history in neuropsychological studies since its original publication in 1935. Originally developed to study cognitive interference directly, the appearance of the “Stroop Effect” has remained remarkably consistent over the past 80 years since the test’s publication in many contexts across a broad range of ages, sociodemographic conditions, and general health status [21]. Still, it has remained challenging to explain the precise cognitive source of the Stroop Effect, and specific processing models are still debated. Many extensive in-depth reviews of the ST and its scoring methods reveal little to no consensus on any cognitive construct that can independently explain all observations in ST performance [18, 22, 23, 16, 21]. Despite this academic debate, it is well-accepted that several cognitive mechanisms are relevant to performance on the test, including speed of processing, automaticity of processing, attention, verbal fluency, reading skills, working memory, and interference/inhibition control. Likely some or all of these mechanisms have combined parallel, additive, and interactive influence on ST performance.

This state-of-the-art gives good reason to hypothesize that differing NDs may impact cognitive mechanisms uniquely to reveal distinguishable patterns in ST performance. Completing these tasks requires engaging many complex cognitive and motor neural circuits, which can express unique impairment and compensation strategies. Despite these distinct impairments and compensation strategies, they may culminate in similar completion times and accuracy rates, making standard metrics useless for the detection or differential diagnosis. More precise quantification and analysis of not only test progress but also motor and behavioral patterns throughout the test may aid in detecting and differentiating ND. We aim to show that by incorporating analysis of eye tracking and audio signals, we can detect these impairment and compensation patterns and create an analytic tool to aid clinicians in the identification and study of ND.

## 3. Prior Literature

### 3.1. Eye Tracking

A review by Antoniades et al. [4] considers the detection and differential diagnostic potential of analyzing eye movement, citing opportunities in the study of AD, PD, Frontotemporal Dementia (FTD), Dementia with Lewy Bodies (DLB), and Huntington’s Disease (HD) to name a few. They conclude in PD to expect dysfunctions such as paucity in blinking, hypometria (target undershooting), increased saccade latencies, directional errors, reduced spatial working memory, and other difficulties in saccade initiation, and further conclude in AD to expect dysfunctions such as disorganized visual scanning, hypometric saccades, prolonged saccade latencies hypothesized to be attributed to attentional-circuits rather than motoric sources, and saccadic intrusions during fixations. Another review by Tao et al. [5] also cites analysis potential in AD, PD, Amyotrophic Lateral Sclerosis (ALS), Multiple Sclerosis (MS), and epilepsy. They conclude in PD to expect higher prosaccadic latency and increased dis-inhibitions, and also note the prevalence of cognitive impairment in PD symptom detection and management, and further conclude in AD to expect dysfunctions such as increased intrusive saccades, less accurate prosaccades, a higher number of saccades, more time to fixate on a target alongside decreased fixation duration, and diminished visual curiosity. They also conclude that errors caused by executive dysfunction are more common in PD, while attention deficits occur more often in AD. Still more reviews exist [24, 25, 26] which cite widespread encouraging clinical data for eye movement relevance in PD, AD, and other NDs. Some commonly suggested metrics include impaired convergence, bradykinesia (slowed movement), impaired vertical gaze, and decreased blink rates in PD; “oblique” non-horizontal microsaccades, more frequent saccadic intrusions, and less accurate visual search in AD. In general, however, most studies involving eye tracking and NDs focus on single-event responses or isolated motor tasks. They typically do not investigate broad visual behavior during the execution of an established neuropsychological task such as the ST.

Tracking the eye gaze and visual interaction behaviors of subjects with PD or AD during multi-word STs compared to controls has not yet been considered. Some consideration of overall performance and cognitive scores based on final output (rather than behavioral interaction) find significant differences in completion time between PD and controls [27], while some found little to no difference between PD and controls [28]. One prolonged PD followup study found a significant increase in fixation duration in PD subjects when measured two years apart, which is believed not to be a result of cognitive impairment [**?**]. It may implausible to predict a singular performance across all persons with PD due to inherent disease heterogeneity and L-DOPA mediated temporal variability of motor performance. In AD, ST completion times are predictive of the development of dementia of Alzheimer’s type [29], and the interference effect has been associated with levels of dementia [30]. In general, studies have only reported single metric performance on the ST, like completion time, or have otherwise presented altered versions of the ST that are easier to analyze, such as single-stimulus presentations, or those requiring alternate responses such as a key press or an additional fixation in a region of the screen, which may limit the possible interaction and symptom expression. Instead, we present the ST as it was originally designed, presenting many stimuli at once and allowing natural pacing and free, unstructured verbal responses of subjects in each task. We then analyze multiple relevant metrics and consider which appear to be most associated with disease type and progression.

One study analyzed eye tracking data of PD patients in a task that was mechanistically similar to the types of motor and cognitive interaction we elicit in the ST by asking subjects to read a page of numbers spaced in a grid [31].

Their approach successfully detected visuomotor dysfunction (*ROC* − *AUC* = 0.973) and used machine learning to show good performance in three-way classification of ND, vision loss, and healthy control (81% accuracy).

### 3.2. Speech Processing

Speech and language features enable rapid neurological diagnosis by incorporating salient aspects of dysfunction across different domains, including cognitive, acoustic, and linguistic properties. Several works have focused on automatic extraction and quantification of clinically-relevant features from speech signals (e.g., zero-crossing rate, Mel-Frequency Cepstral Coefficients, phonemic grouping) [32, 33] and speech transcriptions (e.g., lexical richness, parts-of-speech tags) [34, 35]. Acoustic features such as fundamental frequency (F0) variability, and loudness variability, among others, can be used to assess irregularities in the rhythm and timing of speech that often occur in motor and cognitive decay [36, 37]. Neveler et al. [38] found that a narrower range of fundamental frequency (F0) and increased pause rate are characteristic speech markers in non-fluent/agrammatic primary progressive aphasia. Other studies showed that F0 variability tends to be significantly lower for subjects with PD than healthy controls and can significantly increase after patients receive dopaminergic medications [39, 40]. Toth et al. [41] reported statistical differences for several acoustic parameters (e.g., speech tempo, articulation rate, silent pause, hesitation ratio) between subjects diagnosed with mild cognitive impairment (MCI) and CTLs. Conflicting results are reported on monoloudness: some studies have reported a reduced amplitude variability in patients with PD [42]. In contrast, others have not identified any significant difference between PD and CTL groups [43, 44]. Individuals with PD also demonstrated difficulty modulating speech rhythm and timing organization [45, 46]. As previously noted, the inherent variability of symptom severity, disease severity and diurnal motoric status of PD may confound easy prediction of task performance.

Within the context of neuropsychological tests, Favaro et al. [47] analyzed simultaneously verbal responses to the Cookie Theft Picture (CTP) description task [48] and the ST (third task only) from a cohort of participants with AD, PD, and PDM. They showed that features capturing response informativeness, reaction times, vocabulary richness, and syntactic complexity provided separability between AD and healthy controls. Similarly, F0 variability helped differentiate PD from healthy controls, while the number of salient informational units differentiate PDM from healthy controls. Similarly, Iglesias et al. [49] analyzed spoken responses from three distinct tests: a modified version of the ST (MST), a verb naming task (VNT), and a noun naming task (NNT). In that study, they reported that AD participants had significantly greater reaction times and significantly lower response accuracy than the other groups across tests. In addition, PDM participants, compared to CTLs and PD participants, took a significantly longer time to complete the MST and NNT. In contrast, all the group participants with NDs showed significantly lower confidence during the MST.

Most of the aforementioned studies were limited to the analysis of either linguistic or acoustic features derived from a singular neuropsychological test and narrow focus on one ND at a time. To our knowledge, none of the previous studies explicitly focused on a detailed analysis of the spoken responses to the original ST task in combination with the analysis of samples collected from other modalities like eye-tracking. As explained in the following subsection, this study presents interpretable speech-based features intended to characterize the performance of subjects with NDs and CTLs during the execution of the ST task. Moreover, it presents a joint analysis of speech and eye-tracking samples with the intent of analyzing the complementary of these two modalities in phenotyping the different disorders.

### 3.3. Multi-Modal

There has been a recent emergence in multi-modal recording and analysis of patients with NDs performing various target tasks. One study has analyzed eye tracking and speech production during sentence formulation in a limited multi-modal analysis to characterize word coding and sentence formulation [50]. While demonstrating more significant word-finding errors for PD patients, they found no significant difference between the word-by-word planning of PD and control subjects in their relational picture naming task. Another study by Vásquez-Correa et al. [51] analyzed speech, handwriting, and gait using deep learning approaches to classify patients with PD, reaching ROC-AUC scores as high as 0.99 when classifying subjects as either Healthy or PD. Another study analyzed eye movement with motor responses and EEG activity, also seeing successful classification using differential delay analysis with AUC scores as high as 0.8 in PD detection [52]. Similar studies also exist for AD detection, with studies utilizing speech and eye movement and speech data combined with neuropsychological tests [53]. The authors combined performance on picture description, reading, and memory description tasks with machine learning techniques to obtain AUC scores as high as 0.84 [53], offering a similar analysis to what we present here but using different tasks.

Our multimodal analysis is unique in that we prioritize consideration of interpretable features. In so doing, we hope to facilitate the integration of our results directly into a clinical setting. We aim to extract meaningful interpretable behavioral patterns from our multi-modal recordings. For this reason, we explicitly do not implement machine-learning techniques to extract latent information; rather we employ machine learning and signal processing as tools to obtain interpretable biomarkers. We also specifically target features related to cognitive processing (e.g., number of visual visits/revisits) in addition to the motor and coordination metrics more commonly analyzed in literature for a more complete consideration of measurements and their relationship to our neuropsychological testing protocol.

## 4. Cohort Description

This study focuses on the two most common NDs: PD and AD. We consider patients reporting a significant cognitive decline, including patients with diagnosed AD along with those with MCI which are positive on one or more AD biomarkers, such as those related to medical imaging. For the sake of simplicity, this group is named AD group. We also include a collection of NDs that mimic Parkinsonian symptoms, to test our ability to differentiate AD and PD from other NDs with mimicking symptom profiles. We name this group Parkinson’s Disease Mimics (PDM). We include patients initially diagnosed with Possible PD, by consensus diagnostic criteria as a provisional or working diagnosis but later diagnosed with another ND, including Dementia with Lewy Bodies, Progressive Supranuclear Palsy, Corticobasal Syndrome, Rapid Onset Dystonia Parkinsonism, Gerstmann Schenker Schyuler Syndrome, Essential Tremor, and Wilsons Disease. Including this group for comparison allows us to estimate our ability to differentiate specific symptoms in addition to detecting symptom presence. These groups are all compared with each other and with a group of Healthy Controls (CTL).

We characterize disease symptom severity using two commonly performed clinical tests of cognitive and motor impairment. We use the Montreal Cognitive Assessment (MoCA) [54] to assess cognitive function related to AD and PD. While we also collected the Clinical Dementia Rating scale for these patients, most subjects were in the early progressing stages of the disease with a CDR of 0.5. A handful of subjects had a CDR of 1 (*n* = 3) and 2 (*n* = 2). However, this did not provide a representative enough sample for either rating level to draw meaningful statistical conclusions, and therefore we do not analyze our correlation with this rating. For PD, we utilize the MDS-Unified Parkinson’s Disease Rating Scale (UPDRS) [55] to asses PD symptom severity and specifically separate section three (UPDRS-III) as a measurement of the motor symptoms of our PD patients. We collected MoCA scores for all participants and UPDRS-III scores for subjects with PD.

## 5. Results

This study combines eye tracking and speech analysis to more precisely characterize the performance and impairment during the ST neuro-psychological test. We aim to reveal rich diagnostic and differentiation potential. We extract eye movement and speech characteristics according to the methods described in our Methods section and then combine the extracted features for analysis. We implement all of this analysis automatically, obtaining all our results using an automated pipeline ^2^ to analyze and visualize performance metrics without significant burden to subjects or test administrators. We first extract and consider the analysis potential of existing traditional ST scoring techniques and analyze their relevance to ND characterization and differentiation. We then consider several proposed extracted measurements and validate their relevance to ND by showing a strong correlation with the existing clinical rating scales, including MoCA for cognitive impairment and UPDRS-III for motor impairment. We then present new metrics and metric combinations which reveal more precise performance characterization, demonstrating statistically significant differences between PD, AD, PDM, and CTL.

Differentiating PD, AD, PDM, and CTL groups must be done carefully as there are many confounding factors, the most significant being age. Relevant symptoms of ND often advance simultaneously with both disease progression and increasing age. Further, in our cohort, while PD and AD have similar age distributions (see Table 1), PDM subjects are often diagnosed at much younger ages making their age distributions significantly different. For this reason, we recorded a broad range of ages of CTLs and stratified them to create two control groups, one age-matched to AD and PD subjects and another age-matched to the PDM subjects. These groups are described in detail in the Methods Section, and the appropriate control group is used in statistical comparisons to ensure that age does not confound any differences in distribution.

**Table 1:** Demographics of each subject group.

| Groups | Total | Age<br>(mean, stdev) | Age<br>(min-max) | Female<br>(%) | MoCA<br>(mean,stdev) | UPDRS-III<br>(mean,stdev) |
| --- | --- | --- | --- | --- | --- | --- |
| PD | 30 | 66.4 , 8.9 | 48-79 | 50 | 26.1 , 2.3 | 23.1 , 15.2 |
| PDM | 15 | 61.1 , 15.8 | 27-77 | 27 | 24.5 , 3.3 | 25.0 , 11.5 |
| AD | 26 | 69.2 , 8.9 | 58-84 | 15 | 20.3 , 6.2 | - |
| CTL | 46 | 66.1 , 11.9 | 26-84 | 54 | 27.6 , 1.6 | - |
| CTL-A | 30 | 69.1 , 7.7 | 49-84 | 50 | 27.5 , 1.6 | - |
| CTL-B | 22 | 62.5 , 15.5 | 26-78 | 36 | 27.6 , 1.5 | - |

**Table 2:** Subject Counts after exclusion criteria.

| Task | CTL-A | CTL-B | PD | PDM | AD |
| --- | --- | --- | --- | --- | --- |
| Word-Color Naming Total | 21 | 12 | 24 | 11 | 14 |
| Color-Naming Total | 24 | 13 | 22 | 11 | 16 |
| Word-Naming Total | 23 | 12 | 23 | 11 | 15 |
| Expected Total | 25 | 14 | 29 | 14 | 18 |

### 5.1. Correlation Analysis

We will present results obtained within the correlation analysis performed between the values of the biomarkers and the clinical scores: MoCA for AD subjects and UPDRS-III for PD subjects, to consider the relevance of our extracted metrics with ND severity. We also will consider MoCA in our baseline metrics for PD subjects. A full list of extracted metrics, along with Spearman’s correlation coefficients and *p*-values, can be found in the Appendix. We include in figures 3,4, 5 and 6 a small selection of metrics from individual tasks which have statistically significant correlations with both MoCA and UPDRS. The variety of metrics illustrates the comprehensive nature of our analysis, which includes multiple independent modalities such as saccade characteristics, and number of visual visits to each word throughout the task, blink rates, and gaze position that can all be related to clinically meaningful benchmarks.

#### 5.1.1. Classic Stroop Metrics

First, we extract completion times and analyze them using classical methods to verify the Stroop effect and consider the relevance of traditional ST measures with ND severity. As recommended by Jensen et al. [17], we consider in each of our experimental groups the three recurring phenomena which account for most of the variance in the ST: reading time, color naming difficulty, and interference management. They are found according to Equations 1,2 and 3.

Let *A* = color-naming time, *B* = word-naming time, and *C* = word-color-naming time

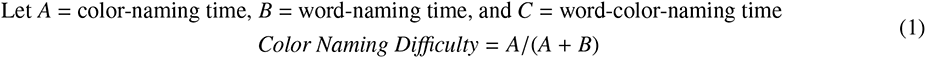

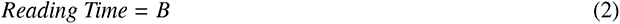

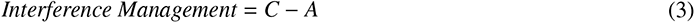

The presence of the Stroop effect in our cohort is easily verified in Figure 2 through the presence of typical values, including positive values for interference management. While most of the regression lines of these classic metrics with disease severity are flat, there are some statistically significant correlations, including increasing Reading Time in AD subjects with worsening MoCA scores (*p <* 0.001) and declining Interference Management PD subjects with worsening UPDRS-III scores (*p* = 0.02), as shown in Figure 2.

**Figure 2:**
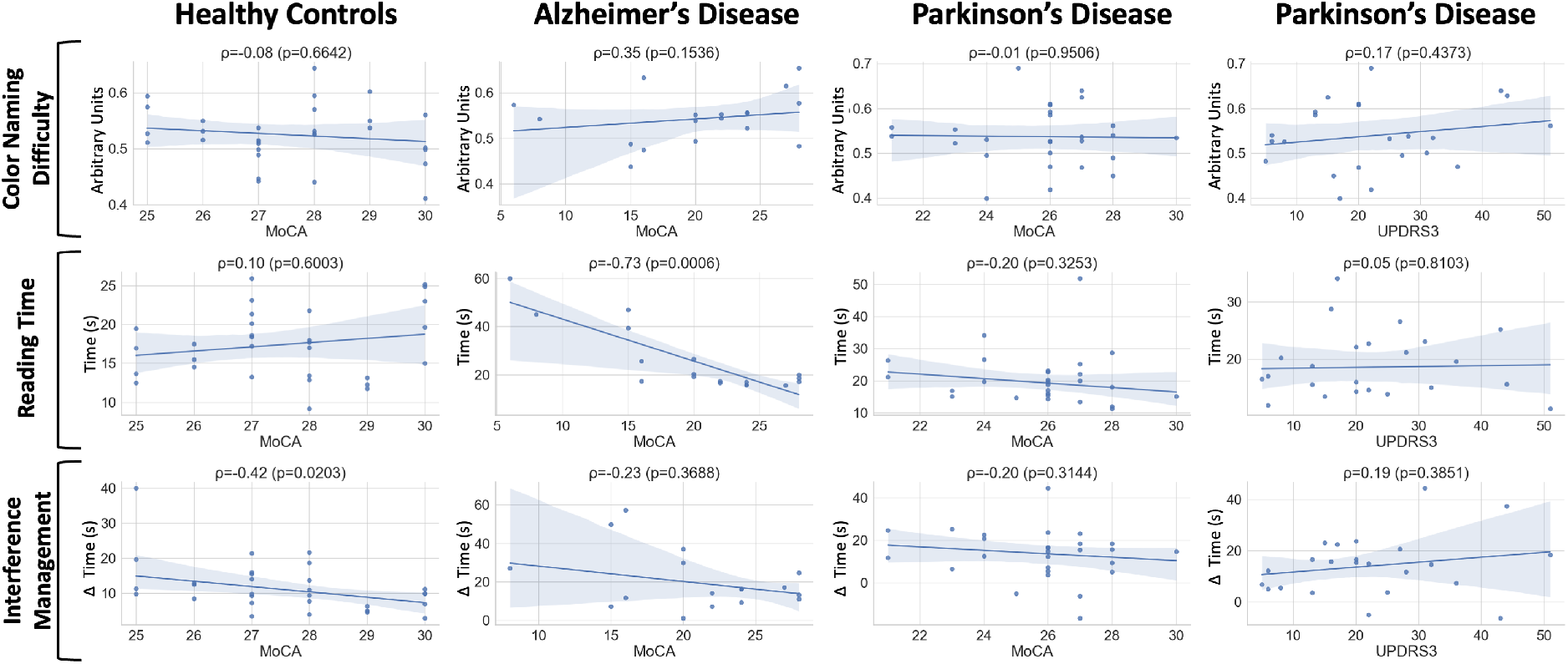
Correlations between classic ST metrics and MoCA scores for all groups. Correlations with UPDRS-III scores are also shown for PD subjects. Spearman’s Correlation Coefficients (*ρ*) and *p*-values (*p*) are reported at the top of each plot. A linear trendline is superimposed on the data, along with a 95% Confidence Interval of the trendline colored in light blue.

Aside from these two metrics, there are no other statistically significant relationships between classic ST metrics and the disease severity of AD and PD subjects.

#### 5.1.2. Eye Tracking

*Single Trial Analysis.* In the word-naming task, we find that AD patients with lower MoCA scores exhibit more saccades of shorter distances and less variability. AD patients with lower MoCA scores fixate on each word for a longer amount of time, revisit those words more often, and have greater variability in their gaze placement when viewing the word. In our case a revisit occurs when subjects shift their gaze forwards/backwards to a new word, and later decide to visually ”revisit” a word they have already looked at. We find that PD patients with worsening UPDRS-III scores tend to blink less often and have more variability in their gaze position when viewing each word. They also tend to move between words with a greater saccade velocity.

In the color-naming task, we find that AD patients with lower MoCA scores again must fixate on each colored patch for a longer amount of time and revisit those colored patches more often and more inconsistently. They also fixate more individual times on a single stimulus and have greater variability in their gaze placement. AD patients with lower MoCA scores spend more time blinking when viewing each color; this correlation holds when normalized for the increased time spend completing the task. We find that PD patients with higher UPDRS-III scores tend to have longer individual blinks, and also revisit the colored patches more often.

In the word-color-naming task, we find that PD patients with worse UPDRS-III scores utilize saccades of a shorter distance with less variability. They must revisit words more often and have a greater standard deviation in the amount of time they view each word. Regarding AD patients, this analysis on this task was excluded as many patients with the most extreme MoCA scores did not finish this task, causing the population of MoCA scores to be unbalanced and therefore exhibit irregular trendlines. However, the lack of ability to finish this task alone may be an indicator of disease severity.

*Combined Trial Analysis.* When comparing performance between the different tasks of the ST using the same relationships captured in Equation 3, we can find more metrics showing a high correlation with rating scales. In addition to those defined relationships, we also considered the simple difference between performance in each task, with results shown also in Figures 3 and 4 for AD and PD, respectively.

**Figure 3:**
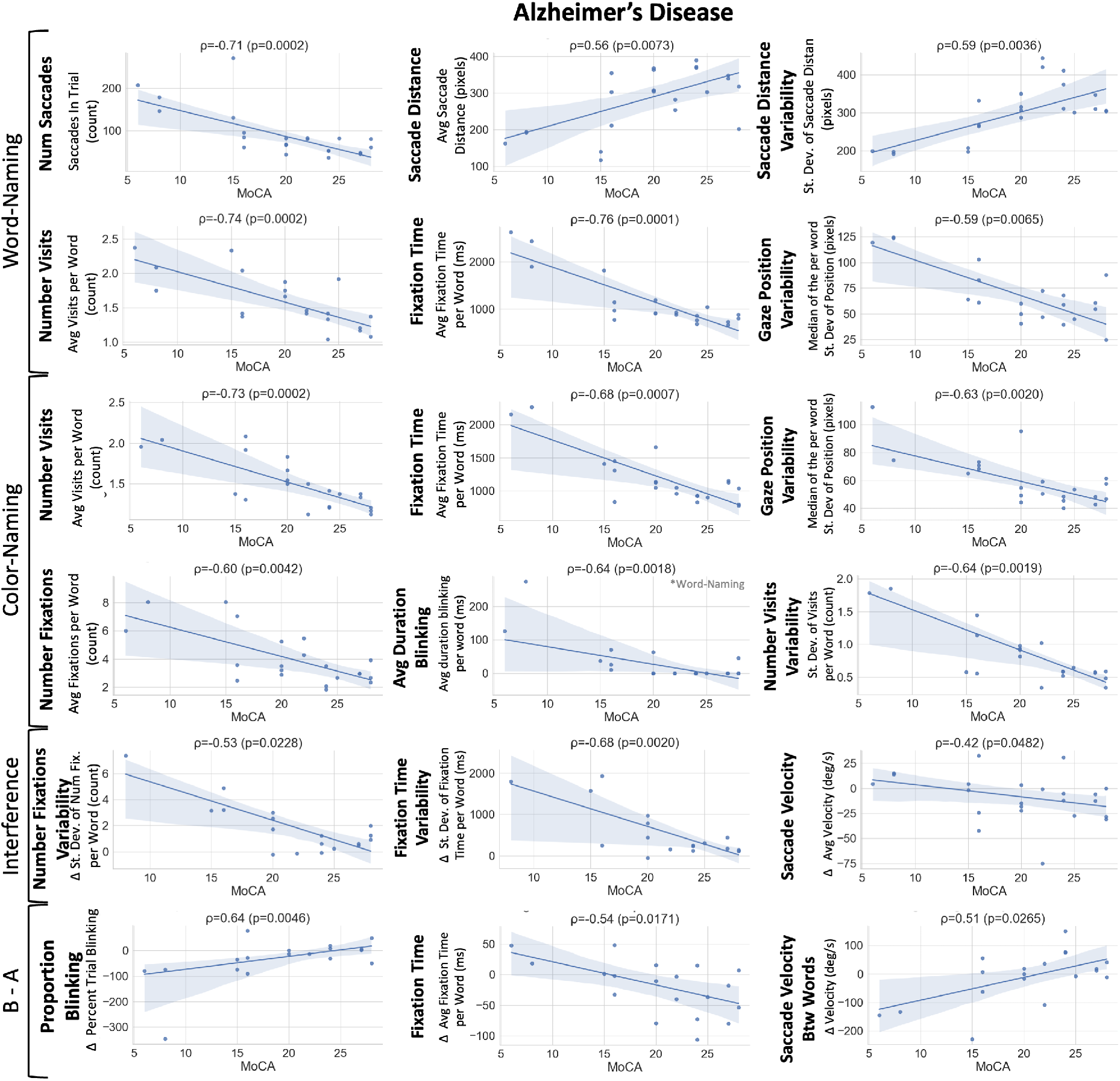
Correlations between eyetracking analysis of AD patients and their associated MoCA scores, used to quantify cognitive ability. Spearman’s Correlation Coefficients (*ρ*) and *p*-values (*p*) are reported at the top of each plot. A linear trendline is superimposed on the data, along with a 95% Confidence Interval of the trendline colored in light blue.

**Figure 4:**
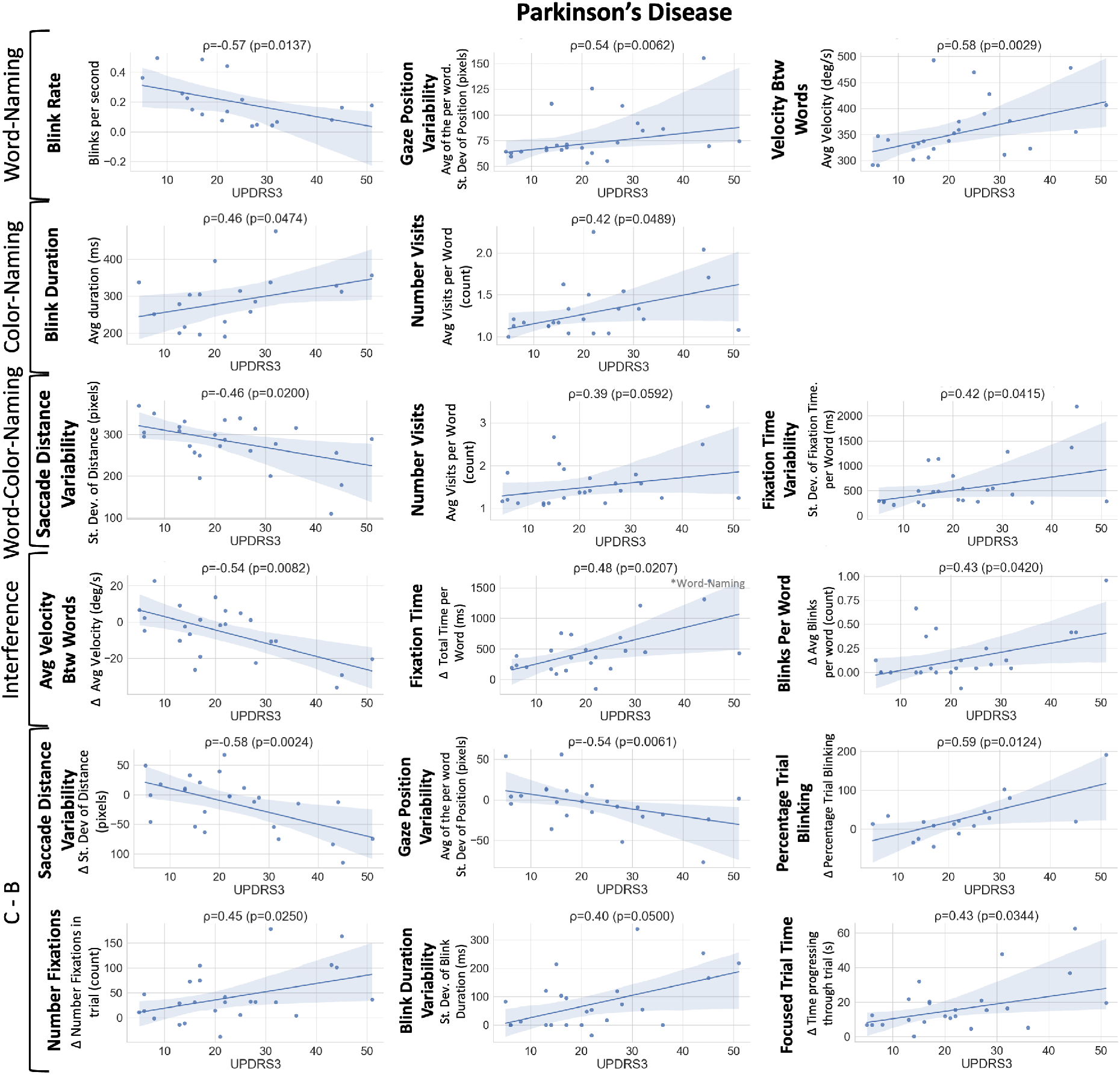
Correlations between eyetracking analysis of PD patients and their associated UPDRS-III scores, used to quantify motor impairment. Spearman’s Correlation Coefficients (*ρ*) and *p*-values (*p*) are reported at the top of each plot. A linear trendline is superimposed on the data, along with a 95% Confidence Interval of the trendline colored in light blue.

**Figure 5:**
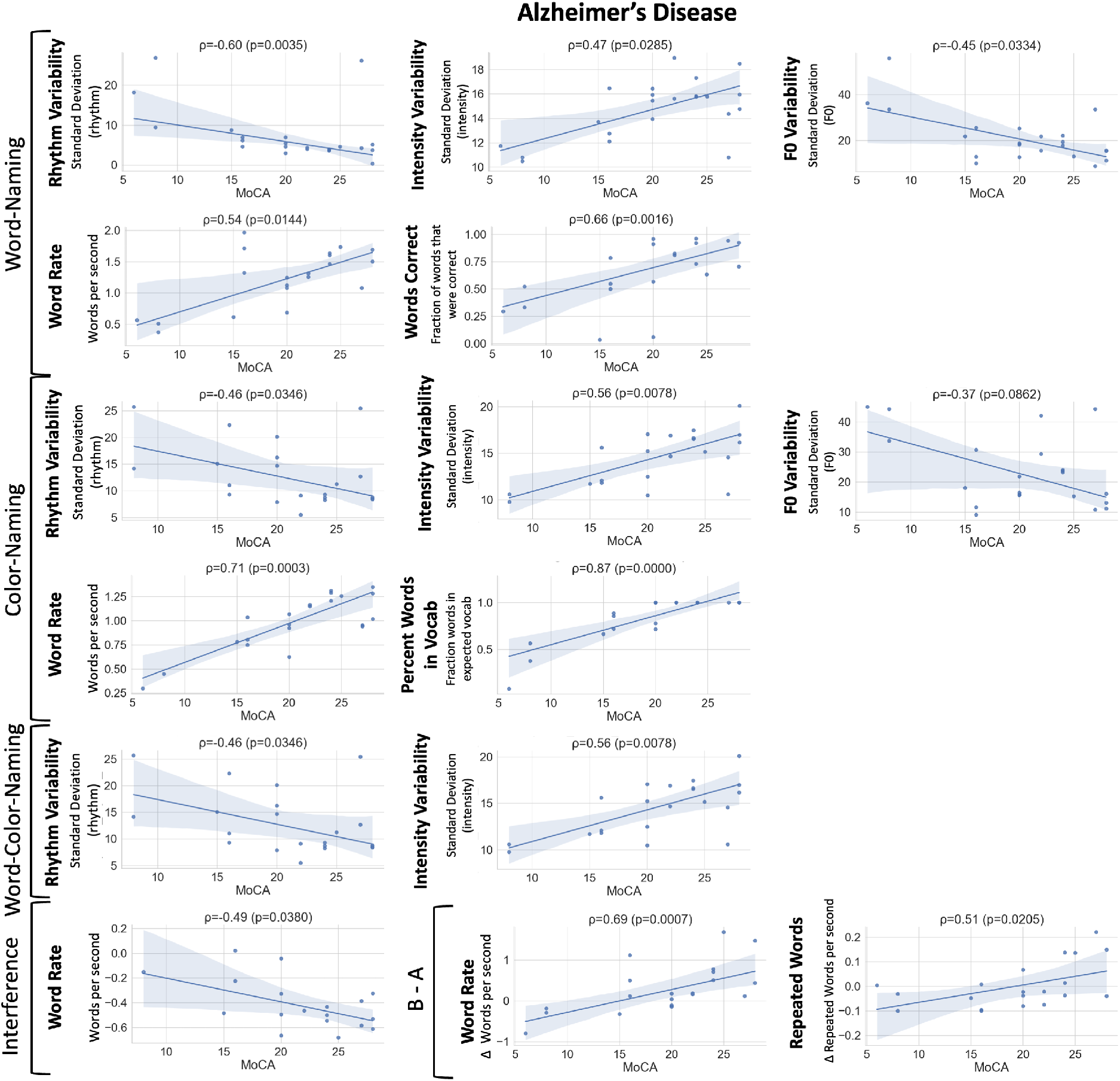
Correlations between speech analysis of AD patients and their associated MoCA scores, used to quantify cognitive ability. Spearman’s Correlation Coefficients (*ρ*) and *p*-values (*p*) are reported at the top of each plot. A linear trendline is superimposed on the data, along with a 95% Confidence Interval of the trendline colored in light blue.

**Figure 6:**
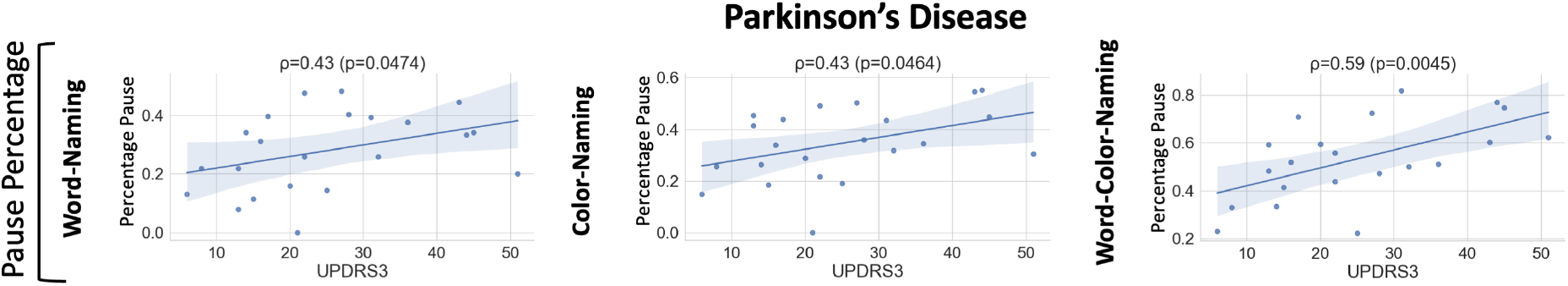
Correlations between speech analysis of PD patients and their associated UPDRS-III scores, used to quantify motor impairment. Spearman’s Correlation Coefficients (*ρ*) and *p*-values (*p*) are reported at the top of each plot. A linear trend-line is superimposed on the data, along with a 95% Confidence Interval of the trend-line colored in light blue.

When quantifying interference management, we find that AD patients with lower MoCA scores tend to manage their interference using longer times looking at each word, and also exhibit a more inconsistent number of fixations along with higher saccade velocities. We find that PD patients with worsening UPDRS-III scores tend to manage interference using lower saccade velocities, and also utilize greater fixation times per word. They also blink more when viewing each word.

When comparing the word-naming and color-naming tasks, we find that AD patients with worsening MoCA scores tend to blink more in the color-naming trial, fixate for longer and move with greater velocity in the word-naming trial. When comparing the word-color-naming and word-naming tasks, we find that PD patients with worsening UPDRS-III scores have a smaller increase in variability in their saccade distances and gaze positions. They do, however see a greater increase in blinking and blink duration variability and also tend to fixate more often in the word-color-naming task. They also see a greater increase in time focusing/progressing through the trial.

#### 5.1.3. Speech

*Combined Trial Analysis.* In the word-naming task, we find that AD patients with lower MoCA scores have significantly greater rhythm standard deviation and higher fundamental frequency variability, but lower intensity variability. They also speak at a lower rate and utter more incorrect responses, accounting for a large number of incorrect spoken responses for the whole trial. These patterns held consistent in the color-naming task, with the additional finding that many of the incorrect words spoken were not in the expected vocabulary of “Red”, “Green”, and “Blue” for this trial.

Instead, there were many other words, often interjections such as “Oh my gosh” or “I messed that up”. We find that PD patients with increasing UPDRS-III scores show a much greater percentage of the trial pausing in all trials.

In the word-color naming task, fewer results were significantly correlated to MoCA scores, yet rhythm variability and intensity variability still showed a significantly negative and positive correlation, respectively, with the MoCA scores. We find that PD patients with increasing UPDRS-III scores show a much greater percentage of the trial pausing in all trials, however, the strongest correlation occurred in the word-color-naming task.

*Combined Trial Analysis.* When comparing performance between the different tasks, we find that AD patients with worsening MoCA scores tend to increase their word rate when managing interference. While not shown, we also find that patients with PD with worsening UPDRS-III scores pause for a greater percentage of the trial when managing interference.

When comparing the word-naming and color-naming tasks, we find that AD patients with worsening MoCA scores tend to see a lower word rate and repeated word instances in word-naming compared to color-naming.

#### 5.1.4. Multimodal Metric Analysis

While our analysis is inherently mutlimodal through simultaneous consideration of eyetracking and speech characteristics, we are also able calculate a few metrics which utilize both modalities to quantify the interactions between visual input and oral output that can only be measured and analyzed in our time-synchronized multi-modal recording setup. As shown in Figure 7, in the word-naming task we find that AD patients with worsening MoCA scores show an increase in time looking at a word before saying the answer and spend less time looking at a word after beginning to say it aloud. The variability in both of these metrics increases drastically for AD patients with worsening MoCA scores. The color-naming task also reports the increase in time spent looking at a word before speaking.

**Figure 7:**
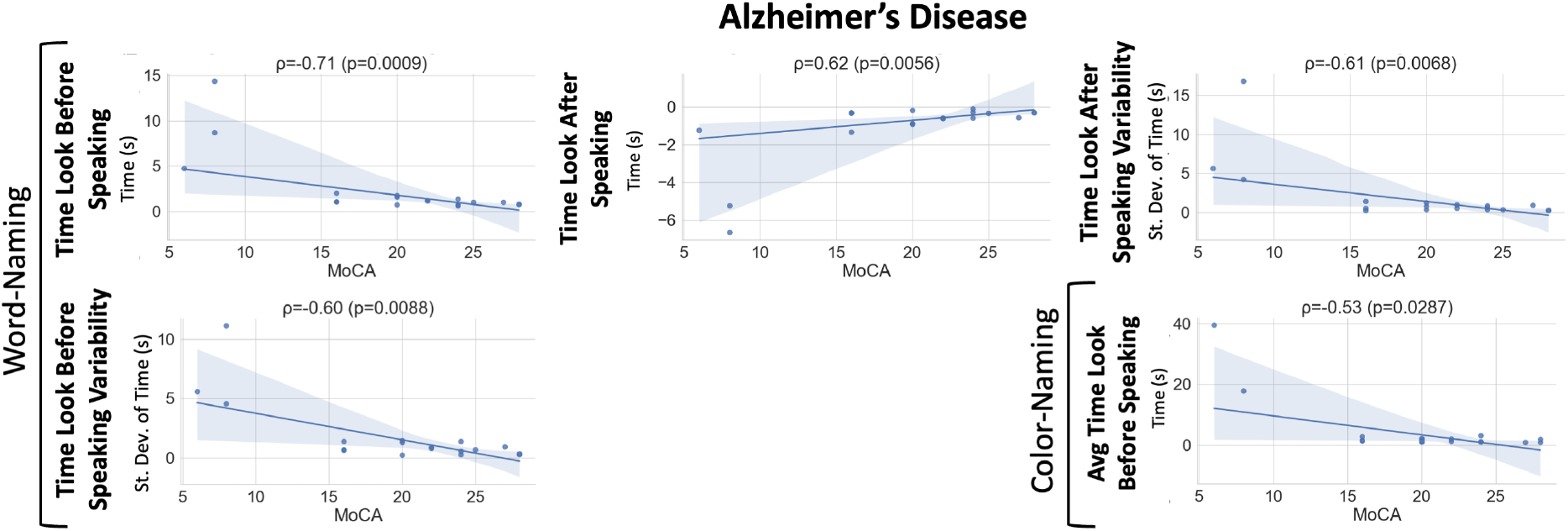
Correlations between multimodal analysis of AD patients and their associated MoCA scores, used to quantify cognitive ability. Spearman’s Correlation Coefficients (*ρ*) and *p*-values (*p*) are reported at the top of each plot. A linear trendline is superimposed on the data, along with a 95% Confidence Interval of the trendline colored in light blue.

### 5.2. Statistical Analysis

We present results obtained in the Kruskal-Wallis H-test comparison, which is a non-parametric version of ANOVA given the observation that not all of our biomarkers are normally distributed, to determine if there were significant differences between the distributions of the different experimental groups for each biomarker in Figures 8 and 9. In the Appendix, for each of the significant biomarkers, we report the corresponding *p*-value, the eta squared effect size (*η*^2^) based on the *H*-statistic and the area under the ROC curve (AUROC). The AUROC of a biomarker can be used as a criterion to measure the biomarker’s discriminative properties. Moreover, we report the observed behavior (OB) of the biomarker that represents the direction in which the median value of the ND biomarker behaved compared to that of the CTL group.

**Figure 8:**
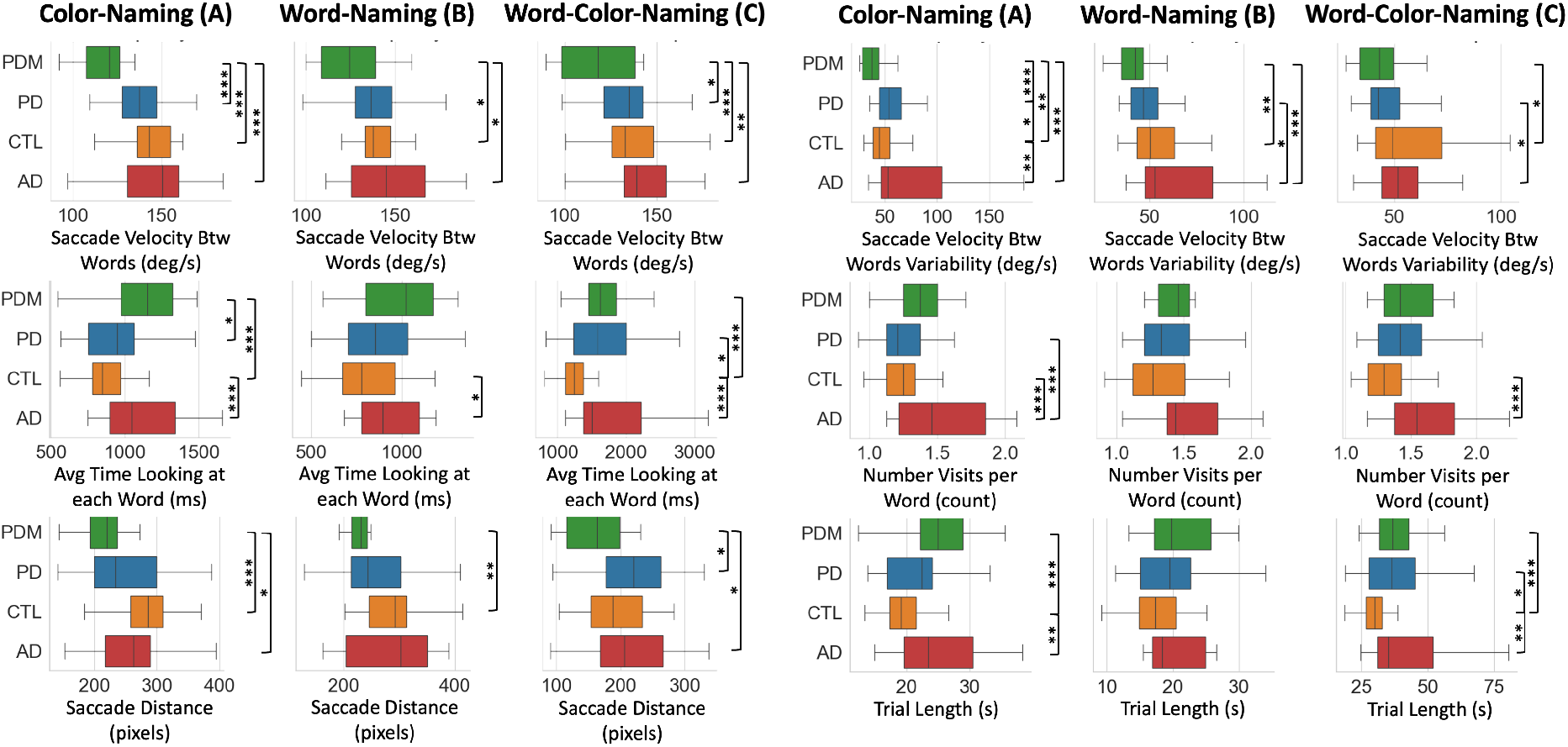
Metrics from eye-tracking analysis demonstrating statistically significantly different distributions between subjects with PDM, PD, CTL, and AD. Each group was compared with a sub-selected age-match CTL group and with each other using a Kruskal-Wallis H-test Test. Statistically significant different distributions are indicated on the right of each graph. * = *p <* 0.05, ** = *p <* 0.02, *** = *p <* 0.01.

**Figure 9:**
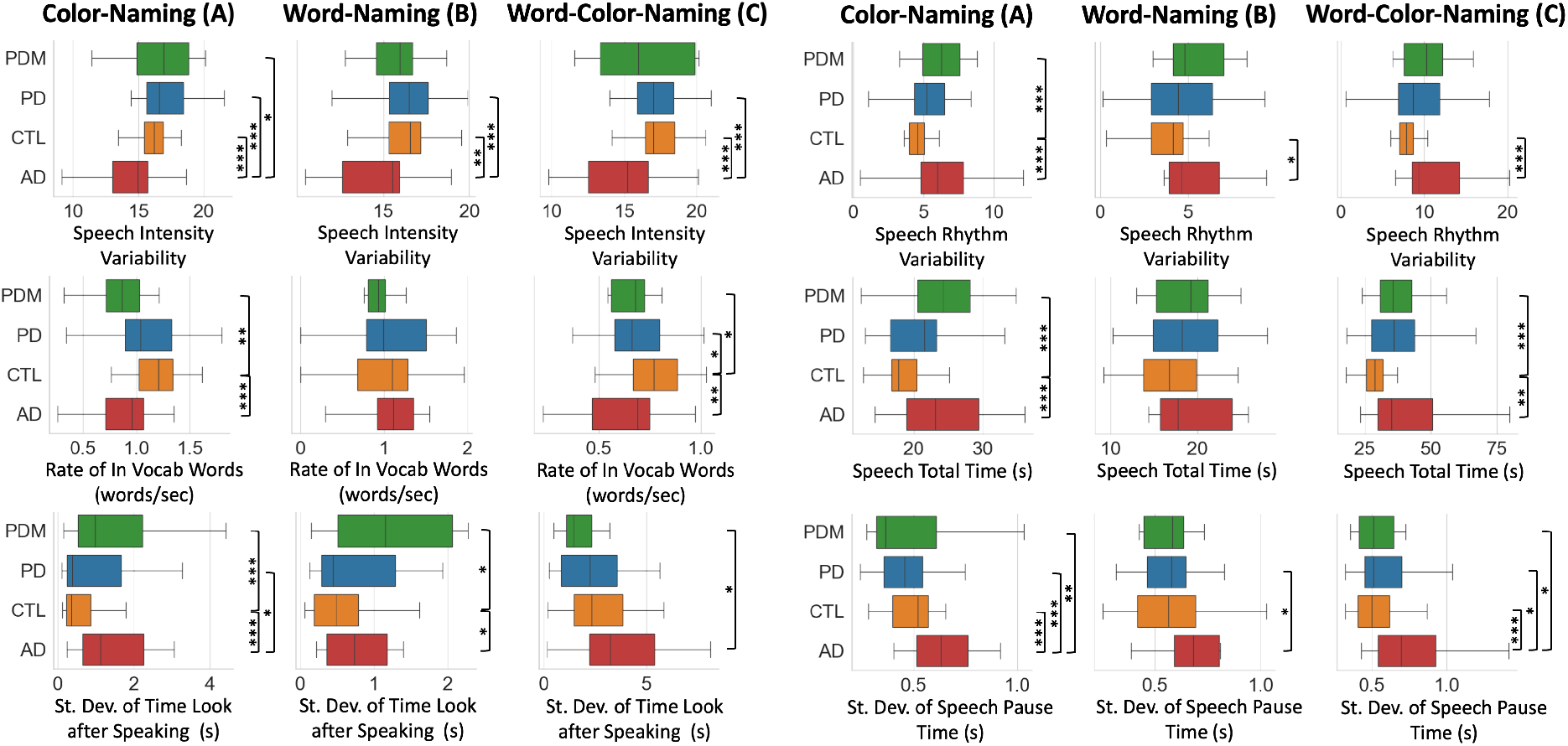
Metrics from individual tasks demonstrating statistically significantly different distributions between subjects with PDM, PD, CTL, and AD. Each group was compared with a sub-selected age-match CTL group and with each other using a Kruskal-Wallis H-test. Statistically significant different distributions are indicated on the right of each graph. * = *p <* 0.05, ** = *p <* 0.02, *** = *p <* 0.01.

*Eye Tracking.* In the color-naming task, compared to CTL patients we find that AD patients spend significantly more time looking at each word, had a greater number of visits than other groups, and also saw greatly increased saccade velocity variability when moving between words. They also saw increased overall trial length. PD patients also saw increased saccade velocity variability compared to CTLs. Most significantly, we find that the PDM groups demonstrate much lower saccade velocity and saccade distance and see greater time-looking and lower saccade velocity variability compared to controls. Note this saccade velocity variability reduction is opposite to the increase in variability that we see in AD and PD patients.

In the word-naming task, we find that AD patients again show increased time looking at each word and increased saccade velocity variability compared to CTL subjects. We find that PD subjects show lower saccade velocity variability compared to AD subjects.

In the word-color-naming task, AD subjects showed increased time looking at each word, number of visual visits, and trial lengths compared to controls. Moreover, AD subjects again exhibited greater saccade velocity variability when compared to PD subjects. Compared to the CTL group, patients with PD also showed increased time looking at each word and also exhibited increased trial length times. Lastly, patients with PDM exhibited significantly lower saccade velocities compared to all groups, lower saccade distances compared to AD and PD subjects, lower saccade velocity variability compared to CTLs, and increased time looking at each word and trial lengths compared to CTL patients.

*Speech.* In the color-naming task, the AD group showed a significantly lower loudness variability than the CTL, PDM, and PD groups. The AD group also shows significantly higher speech time, pause variability, pause percentage, mean pause length, and speech rhythm standard deviation than CTL and PD participants. The PD group reported a significantly greater pause time than the CTL group. The PDM group had a significantly lower F0 standard deviation than the AD group and a significantly greater speech time than the PD and CTL groups.

In the word-naming task, the AD group showed a significantly lower loudness variability than CTL, PDM, and PD groups and significantly greater pause time, pause percentage, pause variability, mean pause length, pause-speech ratio, and speech rhythm standard deviation than the CTL, and the PD groups. The PDM group reported a significantly lower F0 standard deviation than AD, PD, and CTL.

In the word-color naming task, the AD group showed a significantly lower loudness variability than CTL, PDM, and PD groups and significantly greater pause time, pause percentage, pause variability, mean pause length, and speech rhythm standard deviation than the CTL and PD groups. Similarly, the PD and the PDM groups reported a greater pause time than the CTL group.

## 6. Discussion

Our proposed approach captures subject behaviors from a wide variety of behavioral perspectives, all of which can complement each other to promote more stable and reliable metrics than common single-output clinical test results. We can combine speaking rate, visual interaction, and response accuracy throughout three neuropsychiatric tests to get a better picture of subject behavior and performance. We observe that many different modalities, including speaking characteristics, fixation sequences/timing, saccade velocity/variability, and gaze characteristics hold information relevant to the motor and cognitive impairment. This validates that the observations we are making with our system can quantify behaviors reflective of the clinical rating scales and behaviors clinicians already use to aid in diagnosis, and suggests that these measures can be proxies for the cognitive and motor behavior of participants.

We obtained results that suggest a similar behavior of groups observed in previously published studies, including reduced blinking, gaze position variability, and general saccade imprecision [4], impaired convergence [24] for PD subjects, and disorganized visual scanning, increased saccade counts, decreased individual fixation duration alongside increased cumulative fixation time [4, 5] for subjects with AD.

### 6.1. New Metrics

Our results suggest evidence of different coping strategies between subjects with AD and subjects with PD when faced with interference management in the word-color-naming task. When comparing correlations with their respective metrics, we find that while worsening patients with AD show a decrease in the percentage of answers that are correct, decreased word rate, increased rhythm variability, and decreased intensity variability, worsening patients with PD show opposing trends of an increase in the percentage of answers which are correct, and show no change at all in either word rate, rhythm variability or intensity variability. We also find that in cases of interference management, patients with AD will increase their saccade velocities while PD patients will decrease saccade velocities. Upon further inspection of the decline in the percentage of correct answers in patients with AD, we noticed this was often due to the insertion of exclamatory phrases such as “Oh my gosh” or “I messed that up” before answering again.

This may suggest that while both AD and PD experience difficulties with this task, subjects with PD tend to compensate for worsening symptoms by slowing down and focusing on each stimulus individually to make sure they answer correctly through a “slow and steady” approach. In contrast, subjects with AD tend to compensate by attempting to speed up and answer more quickly, even if their initial answers are wrong, through a “spray and pray” approach. While this evidence is marginal and based mainly on regression trends in subjects at various disease severities, it is interesting nonetheless.

### 6.2. Additional Remarks

We observed that under more strenuous tasks like word-color-naming, most metrics saw reduced separation between groups. We typically find much better separation between groups in the easier or more automatic tasks of word-naming and color-naming. A possible reason is that all groups found the word-color-naming task difficult, ultimately crowding everyone to a similar range of reduced performance that eliminates any statistical separation. However, when presented with a less demanding task, CTLs will not struggle, while subjects with NDs will continue to experience difficulties by comparison. This motivates further investigation of these methods in tests that may be less demanding of subjects, such as simple reading, writing, and picture description tasks.

### 6.3. Limitations

It is worth noting that we extracted fewer significant metrics for the differentiation of PD subjects than AD subjects. While we were hopeful that the introduction of cognitive load and/or the interference effect may have exaggerated some of the motor symptoms commonly seen in PD patients [56], we did not see this effect as significantly as expected. However, we could identify a much more robust differentiation of AD subjects. This could be because the ST is primarily a cognitive ability test, specifically targeting higher-level processing and inhibition control. Since high-level cognitive deficits are typically more dramatic in AD than PD [5], this could explain why we see greater differentiation from CTLs on this task. Deficits faced by PD patients are expected to be expressed more in targeted motor control tests, which are not analyzed here but are included in an extended test battery.

Other minor factors of the stimulus presentation also ended up being relevant in some cases. While not a common occurrence, we noticed that if there was noise in eye-tracking signals, it most often occurred and got worse as participants approached the bottom of the screen. As subjects begin to look downwards, their eyelids tend to close as their eyes move down, which further obstructs the pupil, which reduces the accuracy of the pupil identification and overall eye-tracking algorithm quality. This occurred more often in subjects with narrow eye openings as more of the pupil is hidden. While we labeled these instances and removed them, some simple changes to the study profile could avoid this by putting more of the stimuli closer to the top of the screen to keep subjects from looking down and further closing their eyes, along with adjusting the screen/setup to assure that subjects are looking forward/up throughout the task could improve the data quality.

A combination of neuro-psychological tests might even better characterize patients to allow for more consistent and reliable differentiation of even two NDs like AD and PD with control subjects, and even more tests will be required to reliably differentiate the broad scope of ND. Different tests may include more targeted eye movement tasks, object identification or arithmetic tasks, more unstructured picture description or conversational tasks, or more fundamental reading, writing, or critical thinking tasks. These may expose unique and independent dysfunctions in addition to those observed with the ST. Only when several tasks are independently analyzed and combined can we begin to represent a patient’s full symptom profile to characterize the disease state. This analysis of the ST demonstrates a very encouraging start to quantifying symptom states, and future work will broaden the independent and combined analysis of aspects of subject behavior across several tests.

## 7. Conclusions

There is a clear need for more reliable, comprehensive, quantitative tools to provide doctors and researchers with richer analysis and allow them to benchmark patient performance and track progress over time. The combined analysis of eye tracking and speech during a cognitive test like the ST reveals many valid metrics which can help quantify disease state and severity, and may also be relevant in identifying disease-specific symptom profiles. These metrics can not only help to build diagnostic tools in isolation, but can also be combined with other analyses and available data for targeted symptom profiling to help optimize the diagnostic pathway of ND patients. This approach can further improve and expand the usefullness of all neuro-psychological tests in clinical settings, providing unique insight into characteristic and pathological behaviors and of ND patients.

## 8. Materials and Methods

NeuroLogical Signals (NLS) is a dataset collected by the authors of this study at Johns Hopkins Medicine (JHM). Data have been collected in three distinct modalities: speech, eye-tracking, and handwriting. Samples from the first two modalities have been considered in the current study.

### 8.1. Data Collection

We utilized 101 subject recording sessions, including healthy controls and patients with AD, PD, and PDM seen in the Johns Hopkins Health System. 7 of these sessions were recorded in follow-up sessions from subjects already in our dataset. Inclusion criteria for the participants included English native speakers and literate, good eyesight after correction with contacts or non-bifocal glasses if necessary. ND subjects must have received a diagnosis of Clinically Established PD, a ND which was initially diagnosed with Possible PD as a provisional or working diagnosis yet later reached another diagnosis^3^, or MCI/AD. Table 1 contains the demographic statistics of the different groups. A trained research assistant gave participants specific instructions on how to complete each task before the start of each recording session. This study was approved by the Johns Hopkins Medical Institutional Review Board (JHM IRB). All the participants signed informed consent.

To account for the different age ranges of our disease groups, we stratify our CTL group to form two groups for statistical comparison: CTL-A for comparison with PD and AD groups and CTL-B for comparison with PDM groups. Description of age ranges of all of these groups are shown in Tabled 1, and a full age distribution histogram is included in the Appendix.

Our battery of tests was designed and administered using Experiment Builder as provided by SR-Research, which presented subjects with a battery of motor and cognitive tests, including the ST word-naming, color-naming, and word-color-naming tasks. This test battery is freely available ^4^. It was administered to unrestrained subjects sitting at a table approximately two feet from the screen, using a computer visible to the patient.

### 8.2. Eye Movement

We utilized an Eyelink Portable Duo in head-free-to-move mode to register eye movement and gaze information from unrestrained subjects. At the beginning of the session, a standard 9-point calibration and validation sequence was performed, and the eye with the lowest validation error was used for analysis. Task 1 in the ST involves the presentation of a 6-row by 4-column grid of words, including “Red”, “Green”, or “Blue” using black colored text, and subjects are instructed to read the words aloud, progressing from left to right and top to bottom. The exact spacing of this grid can be referenced in Figure 1. Task 2 involves the presentation of an identically spaced grid of “####” characters using either red, green, or blue colored text, and subjects are instructed to say aloud the color of the characters progressing from left to right and top to bottom. Task 3 involves the presentation of another identically spaced grid of words, including “Red”, “Green” or “Blue” words in either red, green, or blue-colored text. The color of the text never matches the word that is spelled. Subjects are instructed to say aloud the color of the text, not the word spelled, progressing from left to right and top to bottom. Before presenting these three tasks, subjects are given ten single-word examples that simulate the task 3 scenario for training purposes. Further description and analysis of these examples can be found in [49].

Data is exported from the Eyelink Portable Duo software and processed using a custom Python library developed by the authors that is freely available ^5^. For each phase of the ST, eye rotational velocity (*deg/s*) and gaze position (pixels) data from the eye with the lowest validation error are extracted and isolated for independent processing. Acceleration is calculated from the velocity data. Blinks are identified as any range of time that is missing data, and the precise start and stop timestamps are identified as the time when either acceleration or velocity rises above a threshold of 25 *deg/sec* and 3,000 *deg/sec/sec*, respectively. Saccades are identified using an algorithm very similar to the description in the Eyelink 1000 User Manual (Section 4.3) describing the Eyelink software and is discussed in [8]. Briefly, this approach simultaneously thresholds both eye velocity and acceleration, requiring both thresholds to be surpassed to identify a saccade. After detection, the precise start/stop timestamps are adjusted when both values have fallen below the threshold. We add rejection of identified saccades that do not result in a net gaze position change of more than 20 pixels (XX degrees). We use a velocity and acceleration threshold of 25 *deg/s* and 3,000 deg/s^2^, respectively. Fixations are labeled as any time point which is not labeled as a blink or saccade. Smooth Pursuit does not occur during the proposed tests, as all stimuli appear at a fixed location on the screen. Metrics describing each saccade, fixation, and blink (SFB) are calculated. All considered metrics are listed in Tables B.3, B.4, and B.5.

Next, each SFB are assigned to the presented stimuli (either words or #’s with a color). The goal is to find which stimulus (word or symbols) the participants are looking at at a certain time and the SFB associated with them in order to measure time spent on a word, revisits, etc. Due to the disease symptoms of some subjects, high-quality calibrations are not always feasible, causing the gaze locations provided by the Eyelink system sometimes to be distorted from the true gaze target. We find that in these situations, the Eyelink system maintains its precision in the measurement of eye movement, but the accuracy of its mapping of eye position to pixel locations worsens. Since this is unavoidable in our testing context, a unique approach to identifying the gaze location’s relationship to the presented stimuli was developed. A visualization of our process is shown in Figure 10. In all subjects, progression to a new line involving the movement from the right side to the left of the screen is easily identifiable. These “newline” actions are identified as any movement from the right 1/3 of the fixation range to the left 1/3 of the fixation range in less than 2 seconds. Fixations before/after these newline events are labeled with their respective line index. If more than the expected 6 lines (5 newline events) is detected, the line with the shortest duration is merged with the preceding or succeeding line based on the nearest average vertical position, as this often occurs when the subject briefly looks back at a previous line. Once each SFB is assigned a line index, fixations within each identified line index are isolated, and four clusters are identified using a K-means algorithm, weighted by the fixation duration, to estimate the locations of the four words in that line. Fixations are then assigned to the nearest cluster center. This line index and word index allow us to assign each fixation to a unique stimulus. Lastly, adjacent saccades and blinks are labeled with the associated starting and ending stimuli.

**Figure 10:**
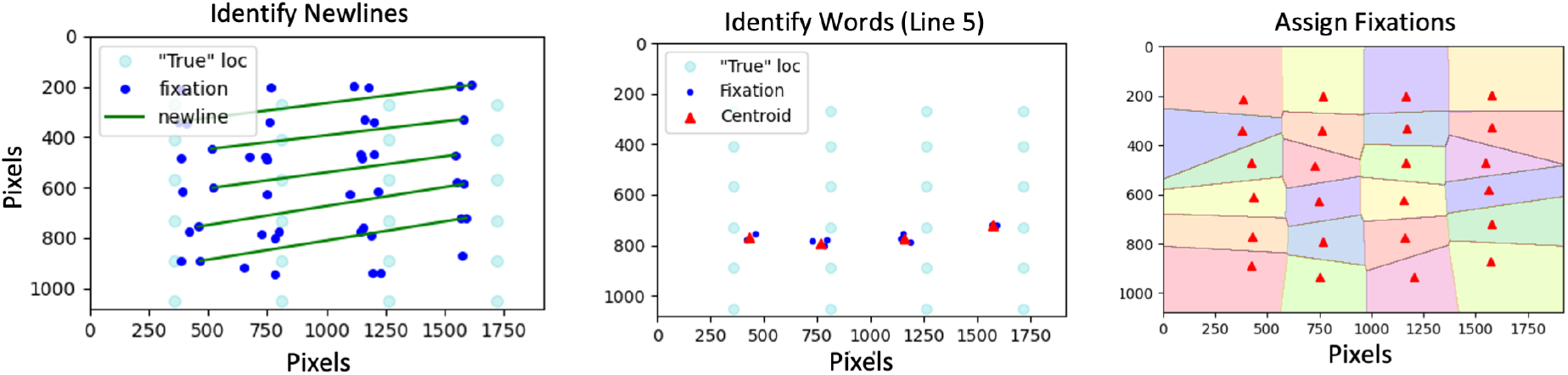
Visualization of the assignment of fixations to stimuli. (LEFT) First “newline” movements are identified as movements from the right 1/3 of the fixation range to the left 1/3 of the fixation range. (MIDDLE) Then each “line” is analyzed individually to identify four clusters representing the words. (RIGHT) Finally, once all stimuli locations have been identified, decision boundaries are drawn, and all fixations are classified.

We validated our approach with visualization and verification of these automatically extracted stimuli assignments in all subjects to ensure the extracted features matched our expected annotations. We show an example of these features and visualizations in Figure 11, verifying our SFB identification and assignment.

**Figure 11:**
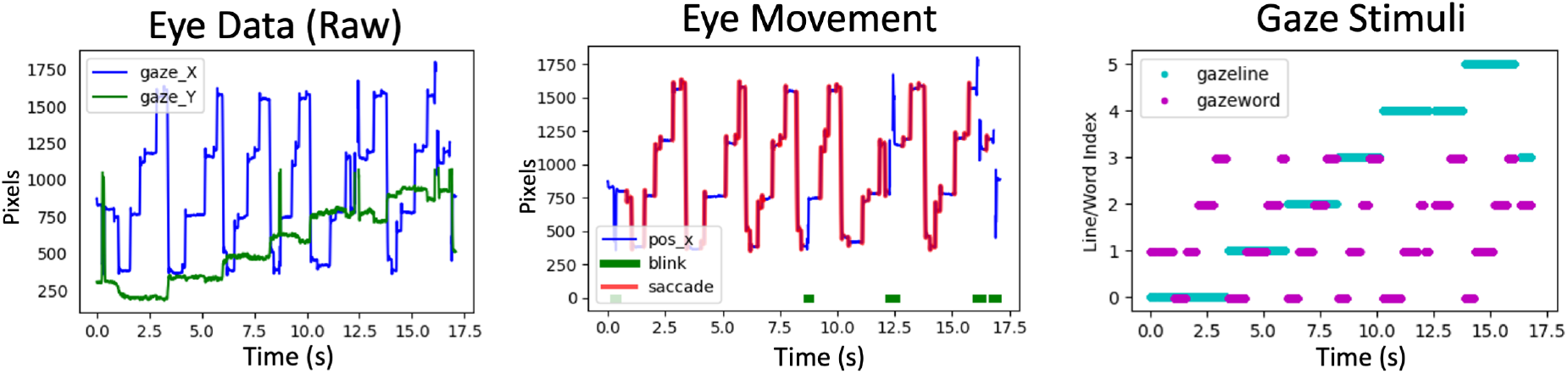
Visualization of the processing of eye movement data. (LEFT) Raw eye movement is extracted. (MIDDLE) Saccades, fixations, and blinks are identified. (RIGHT) Each fixation is assigned to an associated stimulus to indicate where a subject is looking. In cyan, we see the fixations progress from line index 0 through line index 5 throughout the task. Within each line, there are distinctly identifiable fixations on word index 0 through word index 3. Occasionally, as shown in line index 5, the subject looks backward in the stimuli sequence to “revisit” word index 1.

While significant manual data preparation tasks are avoided in favor of autonomous methods of signal cleaning and feature validation, some manual steps were taken to improve the accuracy of this analysis. First, there were many cases where subjects did not promptly begin the task, creating times at the beginning and end of each task with variable fixation progressions. These unordered fixation progressions break from the sequence that is assumed in the stimuli determination algorithm previously described, creating unpredictable SFB word assignments. Therefore, these beginning/ending time ranges had to be labeled by hand and excluded from the stimuli determination functions. After the previously described algorithm identifies stimuli locations using this subset of data, these initially excluded fixations are then assigned to the nearest estimated stimuli location. There were also infrequent occurrences of signal instability, which were primarily due to the presence of glare from glasses or narrowed eye shapes, which affected the identification of the pupil and its exact center, resulting in physiologically improbable data patterns that were apparent outliers. These time ranges were labeled by hand and omitted from the analysis entirely. For each task, only the start and end of the stimuli progression must be identified, and any unstable sections labeled and excluded.

Once all SFB are assigned to a stimulus, summary characteristics describing this subject’s performances culminate. Metrics are generated describing interaction with each word, each line, each column, and each task and saved for later analysis. Plots depicting each step of this process were generated for verification and later reference.

### 8.3. Audio Analysis

*Pre-processing.* An overview of our audio analysis pipeline is shown in Figure 12. Collected recordings, initially sampled at 22 kHz, were re-sampled at 16 kHz as required by the algorithms employed for the feature extraction. The resampling was performed using the sound processing program SoX.^6^ We also applied EBU R128 loudness normalization procedure using the Python library *ffmpeg-normalize*.^7^ This type of normalization leads to a more uniform loudness level compared to simple peak-based normalization. Normalized audios were used to extract the intensity-related feature described below. All recordings were automatically transcribed using OpenAI’s Whisper^8^ [57], an Automatic Speech Recognition (ASR) system trained on 680,000 hours of multilingual and multitask supervised data collected from the web. Moreover, word-starting points were derived using WhisperX [58]. This Python repository improved the accuracy of the timestamps of the Whisper model via forced alignment with phoneme-based ASR models. The phoneme ASR alignment model is language-specific and is automatically imported from torchaudio pipelines^9^. Using the word-starting timestamps, we were able to determine exactly when the subject started delivering their spoken responses and to establish a correspondence between speech and eye-tracking signals. Figure 12 depicts the word alignment process.

**Figure 12:**
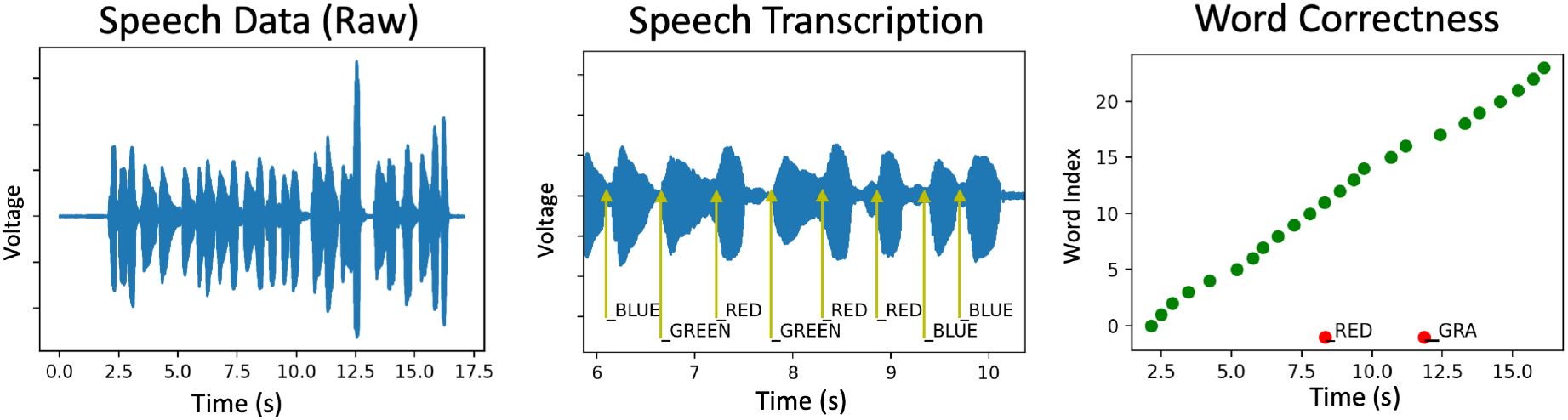
Visualization of the processing of speech data. (LEFT) Raw audio data is extracted. (CENTER) Zoomed in view of the raw data with raw transcriptions and their precise locations annotated. (RIGHT) The sequence of transcribed words is then analyzed to identify if the subjects are uttering words that are correct for the task or incorrect for the task. Green dots indicate a correct utterance in the correct order, while red dots indicate incorrect, out-of-order, or out-of-vocabulary utterances.

**Figure 13:**
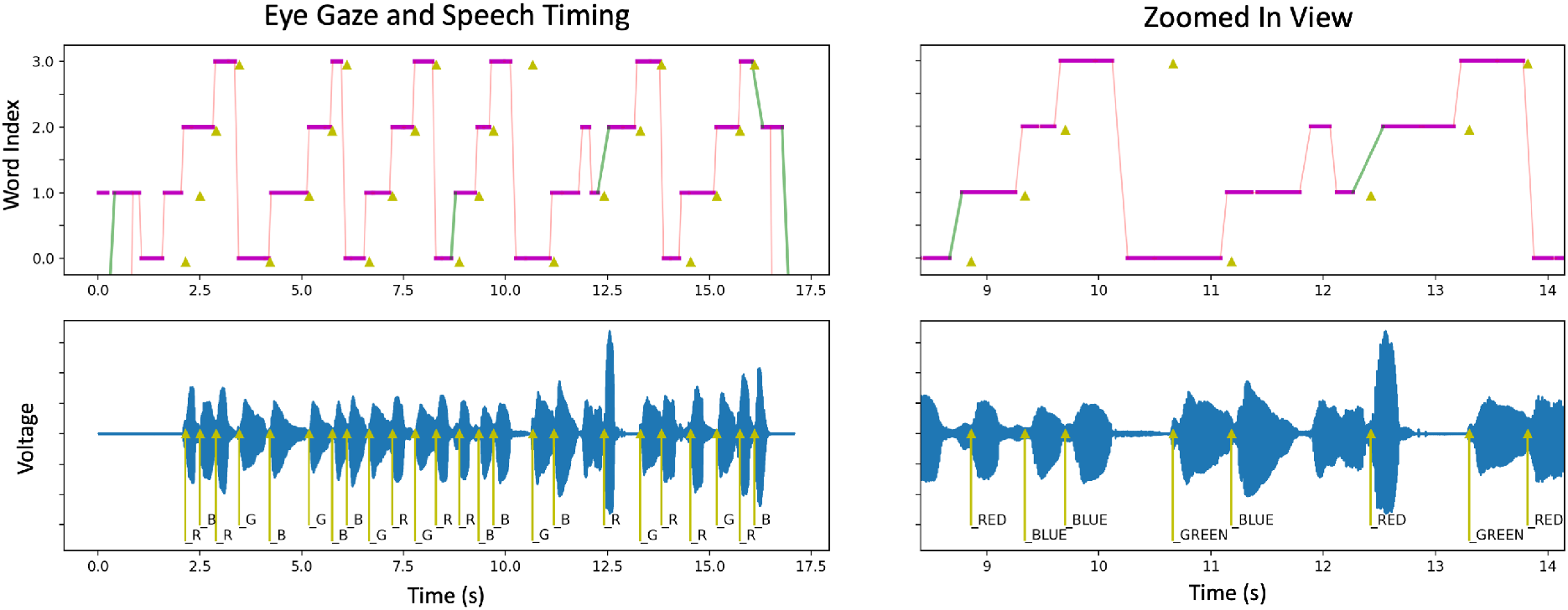
Example of time-aligned eye movement and audio data. (TOP) The visual progression to each word index is shown in pink, with saccades and blinks also shown in red and green, respectively. (BOTTOM) The raw audio data is shown to visualize the time-aligned data. (BOTH) The precise timing of stimuli utterances is annotated in yellow triangles in the top and bottom traces.

*Speech Feature Extraction.* To characterize the spoken production of subjects with NDs and CTLs, we arrange a composite array of features, some already presented in our previous works [47, 49], encoding acoustic and cognitive information. Acoustic features such as fundamental frequency (F0) variability, and loudness variability, among others, can be used to assess irregularities in the rhythm and timing of speech that often occur in motor and cognitive decay [36, 37]. In this respect, it has been shown that patients with right hemisphere (RH) stroke and individuals with selective NDs—e.g., PD, frontotemporal dementia, schizophrenia—may have trouble modulating their tone of voice to express sentence intonation and emotion [59]. We used Parselmouth^10^, a Python library for the Praat software, to compute features such as F0 standard deviation (F0STD) and intensity standard deviation (INTSTD). We also used DigiPsychProsody^11^, an open-access Python repository, to compute features related to speech-time and pause-time such as total speech time, total pause time, percentage pause time, mean pause duration, silence to speech ratio, and pause variability. This library uses the WebRTC Voice Activity Detector^12^. Finally, to model the regularity of speech rhythm (RHYSTD) in terms of the occurrence of the individual words in time. Namely, we measured the time between the starting points of consequent words and computed the standard deviation of these measurements for each recording. To derive the starting point of each word, we computed word alignment using WhisperX.

### 8.4. Multi-modal

Eye Movement and Audio recordings were recorded by the Eyelink system, which allowed us to synchronize the timestamps between the two analyses and consider temporal relationships between eye movement and speech. We use this relationship to help us sort out the correctness of the utterances. First, any out-of-vocabulary words were identified, labeled, and temporarily removed from consideration. Now, left with only in-vocabulary word tokens, we iterate through the ordered stimuli to find the associated utterances. For each stimulus (word), we identify the first timestamp where the subject looks at this stimuli location, find the first matching utterance after this timestamp, and initially assign it to the current stimuli. We then progress to the next word. Suppose the utterance for a particular stimulus is found before the utterance of previous stimuli. In that case, it is assumed that the previously assigned utterance was identified incorrectly (likely belonging to later stimuli), and the previous utterance’s assignment to the previous stimuli index is removed. After iterating through all the expected stimuli, all utterances with an assigned stimuli index are marked as “correct”, and all remaining utterances are marked as “incorrect”. Out-of-vocabulary words are also marked as “incorrect”.

We also considered the relationship of the timestamp of a correct word utterance with the timestamps of the associated visual interaction with this stimuli. We quantify the time looking before utterance as the difference between the first time the subject looks at a word and the beginning of the utterance, and also the time looking “after” the utterance as the difference between the last time the subject looks at a word and the beginning of the utterance. The time looking after the utterance is sometimes negative, as some subjects will look ahead to the next word before beginning to speak. Visualization of this relationship can be seen in figure A.14.

### 8.5. Exclusion Criteria

CTL subjects reporting a MoCA score less than 25 were discarded, as this may be an early indicator of Mild Cognitive Impairment or AD. There were isolated cases of physiologically impossible rapid oscillations between two fixed points observed without the typical acceleration/deceleration profile that is characteristic of saccadic motion, and we considered this data corrupt. This is usually a result of instability in the identification of the pupil and is obvious to identify upon inspection. This occurs more often in subjects with narrowed eyes where the entire pupil is not visible or when there is glare on glass lenses that obstructs the edges of the pupil. Data from a given eye is considered unreliable if more than 20% of data is corrupt or otherwise missing or if all of the expected stimuli focal points are not identified. If data from one eye is considered unreliable, analysis of the other eye is tried, and if these metrics are still not met in the second eye, that subject is discarded from the study. All the audio recordings were supervised to ensure they had appropriate quality. Criteria for acceptable quality included minimal background noise and a task-related response. Corrupted files were discarded from the analysis. When the recordings contained speech from the investigator at the beginning and end, we trimmed the recordings. All valid analysis outputs from relevant tasks are included in our results, even if all three tasks for a given subject are not present or successfully analyzed.

## Data Availability

The authors plan to share all data produced in the present study in the future.

## Acknowledgments

This is a short text to acknowledge the contributions of specific colleagues, institutions, or agencies that aided the efforts of the authors.

## Appendix A. Demographics

**Figure A.14:**
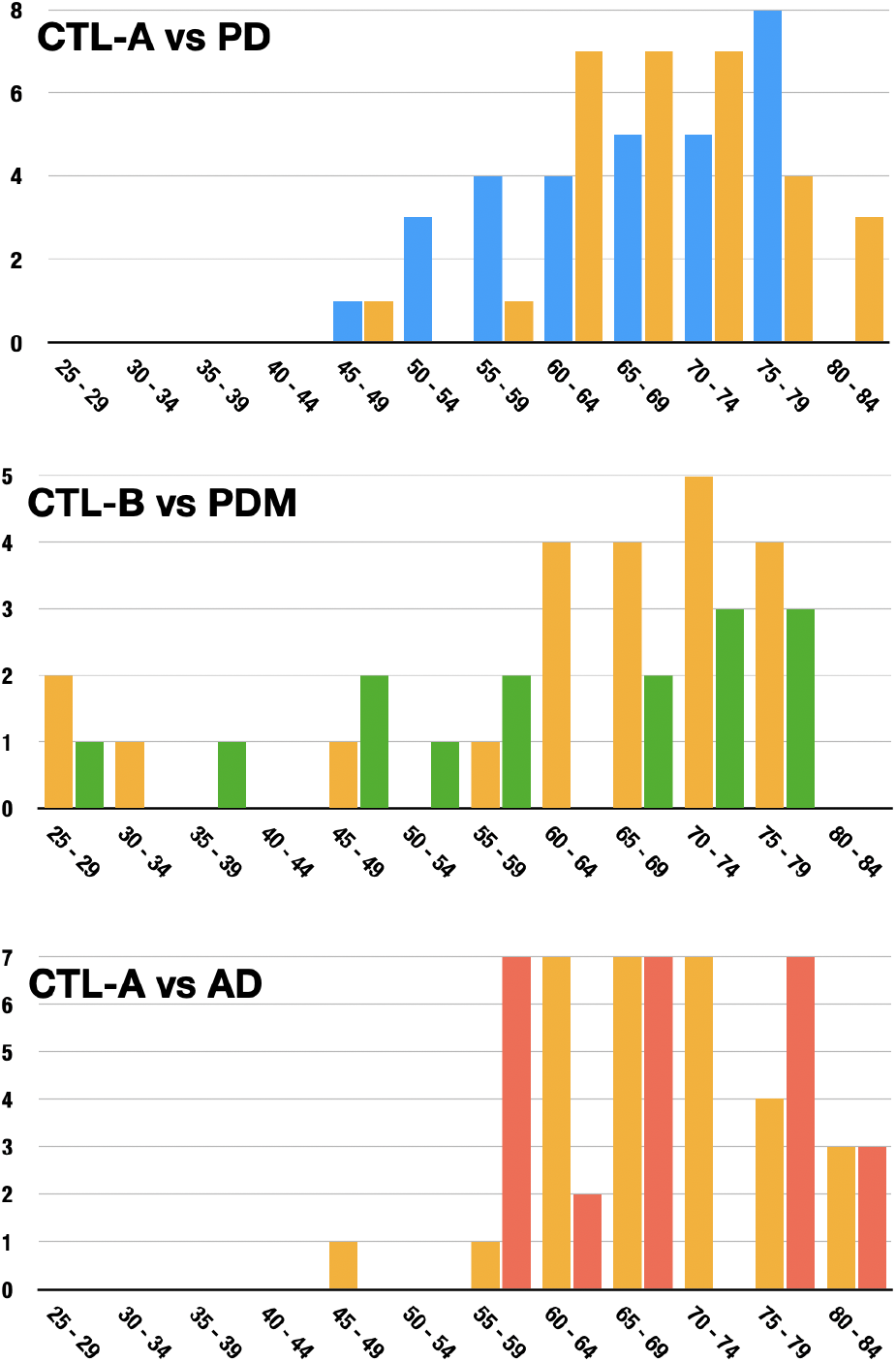
Histograms of the age range of each group and the age-matched control group.

## Appendix B. Calculated Metrics

**Table B.3:** Extracted Metrics – Eyetracking

| Modality | Feature Family | Metric | Units | Source |
| --- | --- | --- | --- | --- |
| Eyetracking | Motor State | Timestamp | ms | NLS Lib |
|  |  | Saccade Position (start & end) | pixels (x,y) |  |
|  |  | Saccade Duration | ms |  |
|  |  | Saccade Distance | pixels |  |
|  |  | Saccade Average Velocity | deg/s |  |
|  |  | Saccade Maximum Velocity | deg/s |  |
|  |  | Saccade Overshoot | pixels |  |
|  |  | Fixation Timestamp | ms |  |
|  |  | Fixation Position | pixels (x,y) |  |
|  |  | Fixation Duration | ms |  |
|  |  | Fixation Number of Blinks | count |  |
|  |  | Fixation Delay since fixation | ms |  |
|  |  | Blink Timestamp | ms |  |
|  |  | Blink Start Position | pixels (x,y) |  |
|  |  | Blink End Position | pixels (x,y) |  |
|  |  | Blink Duration | ms |  |
|  |  | Blink Distance | pixels |  |
|  | Gaze | Number SFB per word | count | NLS Lib |
|  |  | Total Duration SFB per word | ms |  |
|  |  | Average Duration Indiv. SFB per word | ms |  |
|  |  | Fixation Position Std. Dev | pixels |  |
|  |  | Average Saccade Distance | pixels |  |
|  |  | Number Saccades Forward/ Backward/<br>Upward/ Downward (total & per word) | count |  |
|  |  | Number Saccades Forward/ Backward/<br>Upward/ Downward (total & per word) | count |  |
|  |  | Distance Saccades Forward/ Backward/<br>Upward/ Downward (total & per word) | pixels |  |
|  |  | Distance Saccades Forward/ Backward/<br>Upward/ Downward (total & per word) | pixels |  |
|  |  | Saccade Entry and Exit Velocity (avg & max) | deg/s |  |
|  |  | Saccade Entry and Exit Distance (avg & max) | deg/s |  |
|  |  | Number Visits | count |  |
|  | Task Progression | Total Length | ms | NLS Lib |
|  |  | Time before On-Task Progression | ms |  |
|  |  | Number SFB before On-Task Progression | count |  |
|  |  | Time until word 1,2,3... | ms |  |
|  |  | Time until line 1,2,3... | ms |  |

**Table B.4:**
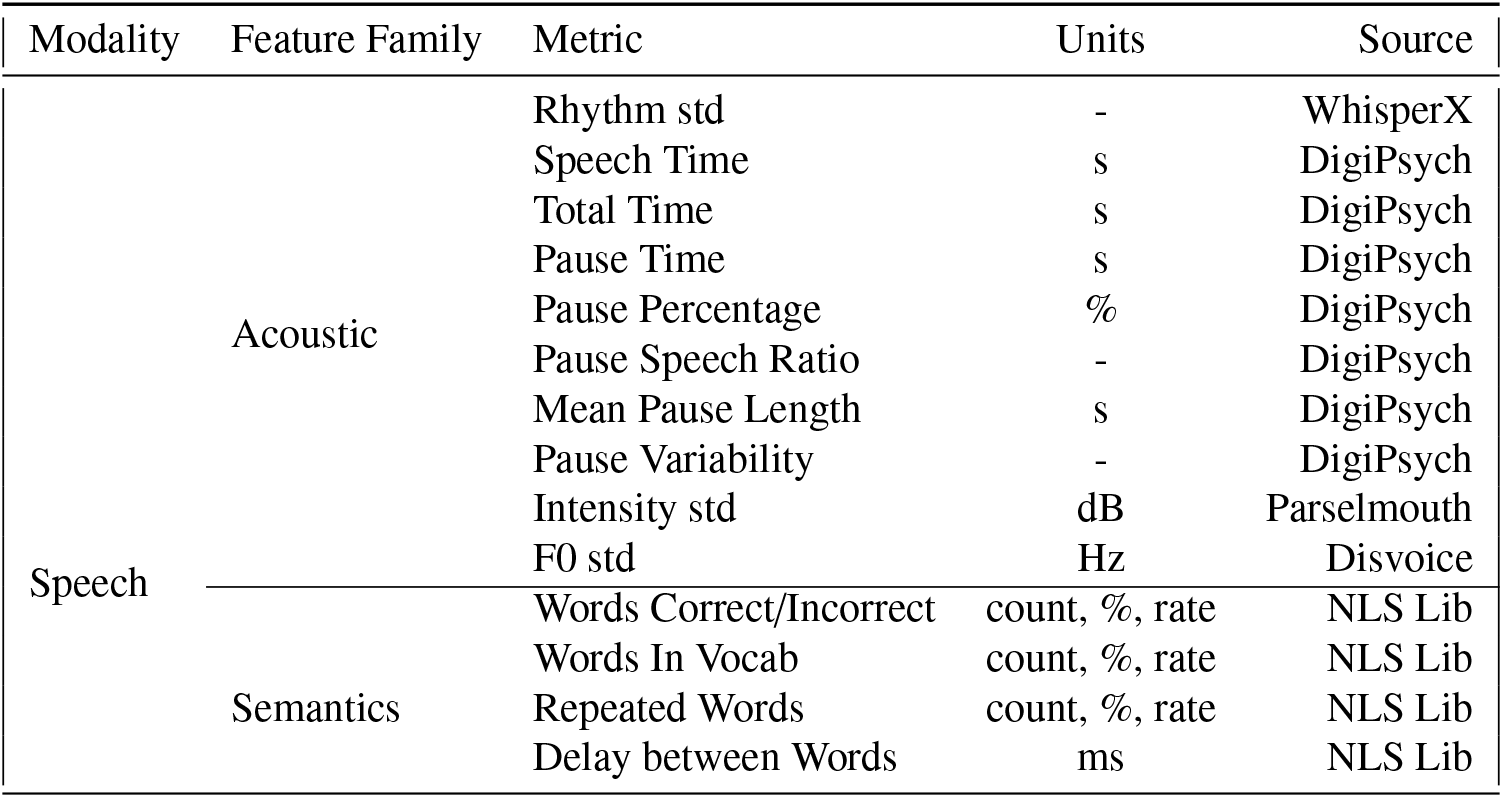
Extracted Metrics - Speech. Abbreviations: std, standard deviation; dB, decibel.

**Table B.5:**
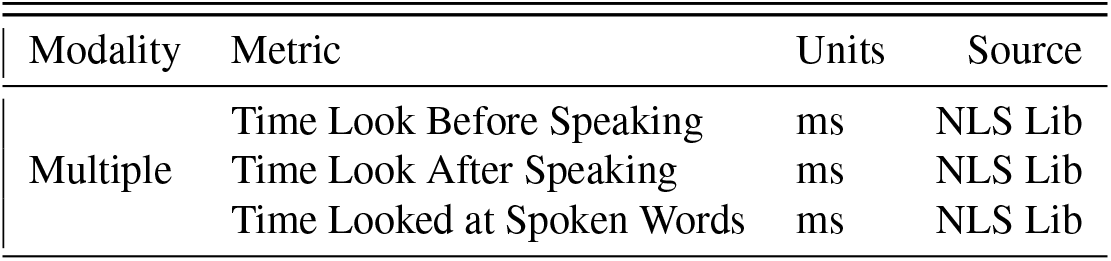
Extracted Metrics - Multimodal

| Modality | Metric | Units | Source |
| --- | --- | --- | --- |
| Multiple | Time Look Before Speaking | ms | NLS Lib |
|  | Time Look After Speaking | ms | NLS Lib |
|  | Time Looked at Spoken Words | ms | NLS Lib |

## Footnotes

1 https://naccdata.org/data-collection/forms-documentation/lbd-31

2 The code for reproducing our experimental pipeline can be found at https://github.com/Neuro-Logical

3 In our study, the PDM group included participants with XXX, YYY, ZZZ…

4 https://github.com/Neuro-Logical/experiment-builder

5 https://github.com/Neuro-Logical/eyetracking

6 https://sox.sourceforge.net/, last accessed on 26 January 2023.

7 https://pypi.org/project/ffmpeg-normalize/, last accessed on 26 January 2023.

8 https://openai.com/blog/whisper/, last accessed on 4 February 2023.

9 https://pytorch.org/audio/main/pipelines.html, last accessed on 26 January 2023.

10 https://parselmouth.readthedocs.io/en/stable/, last accessed on 11 March 2023.

11 https://github.com/NeuroLexDiagnostics/DigiPsych_Prosody, last accessed on 26 January 2023.

12 https://github.com/wiseman/py-webrtcvad, last accessed on 12 March 2023.

